# Predictors of COVID-19 Vaccine Acceptability Among Refugees and Other Migrant Populations: A Systematic Scoping Review

**DOI:** 10.1101/2023.09.15.23295608

**Authors:** Yasaman Yazdani, Poojitha Pai, Shahab Sayfi, Arash Mohammadi, Saber Perdes, Denise Spitzer, Gabriel E. Fabreau, Kevin Pottie

## Abstract

**Objective:** This study aimed to map the existing literature to identify predictors of COVID-19 vaccine acceptability among refugees, immigrants, and other migrant populations.

**Methods:** A systematic search of Medline, Embase, APA PsycInfo and Cumulative Index of Nursing and Allied Health Literature (CINAHL) was conducted up to 31 January 2023 to identify the relevant English peer-reviewed observational studies. Two independent reviewers screened, selected studies, and extracted data.

**Results:** We identified 34 cross-sectional studies, primarily conducted in high income countries (76%). Lower vaccine acceptance was associated with mistrust in the host countries’ government and healthcare system, concerns about the safety and effectiveness of COVID-19 vaccines, limited knowledge of COVID-19 infection and vaccines, lower COVID-19 risk perception, and lower integration level in the host country. Female gender, younger age, lower education level, and being single were associated with lower vaccine acceptance in most studies. Additionally, sources of information about COVID-19 and vaccines and previous history of COVID-19 infection, also influence vaccine acceptance. Vaccine acceptability towards COVID-19 booster doses and various vaccine brands were not adequately studied.

**Conclusions:** Vaccine hesitancy and lack of trust in COVID-19 vaccines became significant public health concerns within migrant populations. These findings may help in providing information for current and future vaccine outreach strategies among migrant populations.

## Introduction

The COVID-19 pandemic poses a significant threat to public health, not only in terms of its impact on mortality and morbidity, but also due to the social and economic disruptions it has caused, as well as the burden of public health restrictions (1). To combat this, one of the most crucial strategies is widespread vaccination among the population (2).

In addition to barriers to equitable access to COVID-19 vaccines, vaccine hesitancy compounds the challenges faced, increasing the difficulties to achieve widespread vaccination coverage. Vaccine hesitancy is defined as the delay or refusal of vaccination despite its availability, by the WHO Strategic Advisory Group of Experts on Immunization (SAGE) working group on vaccine hesitancy (3).

Understanding the factors that contribute to vaccine hesitancy is a complex and context-specific task. For COVID-19 vaccines, skepticism has arisen due to the novelty of the disease and concerns about their safety and efficacy. These concerns have been amplified by the prevalence of false or misleading information, or misinformation, leading to what the WHO has labeled an “infodemic” (4).

Previous research indicates that certain populations, including Refugee, Immigrant, and Migrant (RIM) populations, are at higher risk of vaccine hesitancy (5). While there is limited research on the determinants of vaccine hesitancy in these groups, studies have shown that lack of accessibility, mistrust stemming from culturally insensitive healthcare practices, and discrimination experienced when seeking care are common reasons (6). Furthermore, RIM populations face elevated risks of COVID-19 exposure due to their overrepresentation in high-risk occupations, crowded living conditions, social deprivation, and barriers to accessing reliable information on preventive measures (7). Identifying predictors of COVID-19 vaccine acceptance in RIM populations could inform strategies to overcome potential obstacles and ensure effective vaccination campaigns.

### Research Objectives

We conducted a systematic scoping review using an existing conceptual framework to map the English literature on COVID-19 vaccine acceptability predictors among RIM populations, and to identify the current gaps in the literature.

## Materials and methods

In this review, we applied the enhanced version of the Joanna Briggs Institute (JBI) scoping review guidance (8).

### Ethics statement

N/A

### Eligibility criteria

The International Organization for Migration (IOM) defines a migrant as “A person who moves away from his or her place of usual residence” (9). Our study focused on predictors of COVID-19 vaccine acceptability among international migrants. A list of the relevant terms is available in Table 1.

**Table 1.** Population, concept, and context (PCC) and relevant keywords.

| **PCC elements:** | **Search terms** | **Search results** |
| --- | --- | --- |
| **Population (Refugees and other migrant populations)** | Immigrant, migrant, emigrant, refugee, asylum seeker, diaspora, newcomer, foreigner, foreign-born, foreign worker | 750 |
| **Concept (vaccine acceptability)** | Immunization, vaccine, vaccination, vaccine hesitancy, vaccination hesitancy, vaccine refusal, anti-vaccine, hesitancy, hesitation, trust, acceptance, refusal, willingness, attitude, choice, denial, phobia, avoidance, decision, uptake, doubt, resistance, reluctance, exemption, controversy, dilemma, intention, skeptic, delay, distrust, mistrust, confidence, acceptability, perception, concern, fear, belief, dropout, rejection, behavior, knowledge |  |
| **Context (COVID-19 pandemic and vaccination)** | Covid-19, severe acute respiratory syndrome coronavirus 2, Coronavirus, Sars-cov-2 |  |

We conducted a comprehensive review of all observational, peer-reviewed studies published in English up until January 31, 2023, as it aligns with the scope of our research question. Studies focusing on predictors of COVID-19 vaccine acceptability and similar concepts among RIM were included. We also included general population studies, if more than 50% of their participants were immigrants or refugees, or a separate sub-group analysis of RIM were reported.

Reviews, experimental and qualitative studies, editorials, and commentaries were excluded. However, the references of relevant reviews were screened in order to find any eligible original research articles.

### Search methods

Keywords, and index terms suggested by refugee and immigrant health professionals and the Western University research librarian, helped us develop a comprehensive search strategy for the targeted databases. We searched the following databases, without any search restrictions: Medline, Embase, APA PsycInfo and Cumulative Index of Nursing and Allied Health Literature (CINAHL). We also searched the World Health Organization (WHO), The United Nations High Commissioner for Refugees (UNHCR) and the International Organization for Migration (IOM) websites for relevant information. Our full Medline search strategy is presented in appendix 1.

### Screening and selection

Following the search, all identified publications were collated and uploaded on COVIDENCE (a web-based platform that manages systematic review data) to review and remove duplicates. All the stages of screening were done separately by two reviewers. Any conflicts between primary reviewers were resolved through discussion and getting the opinion of a third senior reviewer.

### Data Extraction and Management

Two reviewers extracted and charted the data, using a data extraction tool based on a previously reported conceptual framework (Appendix 2) (10).

### Synthesis of the results

Finally, we presented the results according to the Preferred Reporting Items for Systematic reviews and Meta-Analyses extension for Scoping Reviews (PRISMA-ScR) Checklist in tabular form, accompanied by a narrative summary. As a scoping review, our study aimed to provide an overview of the existing literature rather than assessing the quality of included studies.

### Results

Using the above-mentioned search criterion, the databases yielded 1635 total articles. No further relevant studies were found on WHO, IOM or UNHCR websites. The details of the study selection process are shown in Figure 1.

**Fig 1.**
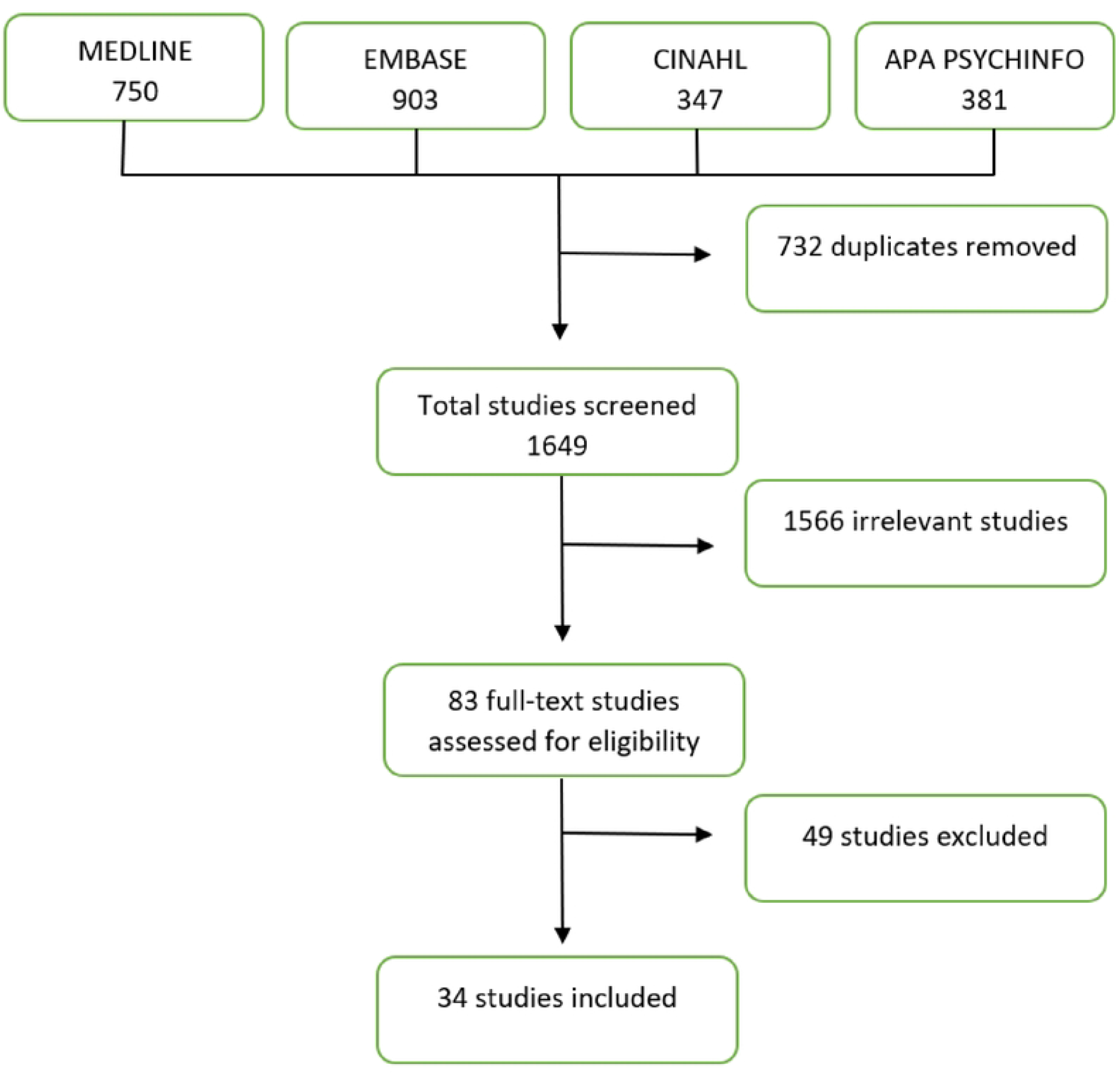
**Selection of included studies (PRISMA Flow diagram)**

### Characteristics of included studies

Thirty-four observational studies were found to meet the inclusion criteria. We present a descriptive analysis of the studies included in Table 2. Most studies were conducted nationally (89%), in high income countries (76%). Forty-eight percent of all migrant participants were female and 52% were male. Eight (24%) and two (6%) articles studied refugees and undocumented migrants respectively, while only one (3%) focused on international students exclusively. Only two studies (6%) examined migrants’ hesitancy towards getting COVID-19 booster dose and three (9%) evaluated participants’ preferences regarding different vaccine brands offered or their country of origin. Ninety percent of studies were conducted after the first WHO recommendation regarding COVID-19 vaccination, released in December 2020.

**Table 2.** Characteristics of included studies.

| **Study characteristics** | **Study attributes** | **Number of studies (%)**<br>**N=34** | **Study ID** |
| --- | --- | --- | --- |
| **Host country** | **High income** | 26 (76%) | 12-14,17-24,26-28,30,32,33,35-37,40-42,50,55,63 |
|  | **Middle income** | 7 (21%) | 15,16,25,34,36,38,39 |
|  | **Low income** | 1 (3%) | 31 |
|  | **USA** | 11 (32%) | 17,20,22,24,28,30,32,41,50,55,63 |
|  | **Germany** | 3 (9%) | 13,18,40 |
|  | **South Korea** | 2 (6%) | 12,35 |
|  | **Australia** | 2(6%) | 23,26 |
|  | **Qatar** | 3 (9%) | 14,19,21 |
|  | **Thailand** | 2 (6%) | 25,34 |
|  | **Lebanon** | 2 (6%) | 16,38 |
|  | **other** | 10 (29%) | 26,27,29-31,33,36,37,39,42 |
| **Geographical coverage** | **National** | 30 (89%) | 12-28,31,33-35,37-42,50,55,63 |
|  | **International** | 4 (11%) | 29,30,32,36 |
| **Study design** | **Cross sectional** | 34 (100%) | 12-42,50,55,63 |
|  | **Mixed method** | 2 (6%) | 41,55 |
| **Sampling techniques** | **Convenience sampling** | 12 (35%) | 14,16,21,23,25,26,35,37,39-41,50 |
|  | **Snowball sampling** | 4 (12%) | 12,13,20,22 |
|  | **Probability sampling** | 6 (18%) | 17,19,27,28,34,42 |
|  | **Not mentioned** | 12 (35%) | 15,18,24,29-33,36,38,55,63 |
| **Sample size** | **Range** | (127-7821) |  |
|  | **Less than 500** | 17 (50%) | 13,15,19,20,22,23,28,32,35,36,39-42,50,55,63 |
|  | **500-1000** | 6 (18%) | 12,19,25,26,30,37 |
|  | **More than 1000** | 11 (32%) | 14,16-18,21,24,27,29,31,33,38 |
| **Gender** | **Male** | 20412(52%) |  |
|  | **Female** | 18630 (48%) |  |
| **Other demographic characteristics** | **Racialized status/ethnicity** | 11 (32%) | 12,14,17,19,20,24,32,35,41,50,63 |
|  | **Immigration status** | 16 (47%) | 15,19,22,24,28,30-32,34,36,38-42,63 |
|  | **Region of origin** | 19 (56%) | 13,16,18,22-25,28-30,34,36,38-42,50,55 |
| **Target population** | **Refugees** | 8 (24%) | 15,16,22,26,31,38,39,41 |
|  | **Undocumented migrants** | 2 (6%) | 30,32 |
|  | **Migrant workers** | 2 (6%) | 34,36 |
|  | **International students** | 1 (3%) | 42 |
|  | **Any type of migratory background/status** | 9 (26%) | 12,21,23,28,29,33,35,40,50,55 |
|  | **Migrants as a subgroup of general population** | 16 (47%) | 13-21,24,25,27,29,31,37,63 |
| **Timeline** | **Before WHO first COVID-19 vaccination recommendation** | 4 (12%) | 14,21,27,50 |
|  | **After WHO first COVID-19 vaccination recommendation** | 30 (88%) | 12,13,15-20,22-26,28-42,55,63 |
| **Vaccine outcomes** | **Vaccine acceptance** | 9 (26%) | 12,20,22,25,34,35,37,39,42 |
|  | **Vaccine hesitancy** | 11 (32%) | 15,17,19,21,24,26,27,29-31,36 |
|  | **Vaccine intention** | 14 (42%) | 13,14,16,18,22,23,28,32,33,38,41,50,55,63 |
|  | **Hesitancy towards booster dose** | 2 (6%) | 23,24 |
|  | **Acceptability based on vaccine brand/country of origin** | 3 (9%) | 16,25,39 |

Table 3 shows the variables examined in the included studies; however, only some studies investigated the association of these factors with vaccine acceptance/intention/hesitancy (Table 4). According to the literature review and based on the conceptual framework used (10), we categorized predictors of COVID-19 vaccine hesitancy into four major groups.

**Table 3.** Factors examined in included studies.

| **Study factors** | **Number (%)**<br>**N=34** | **Study ID** |
| --- | --- | --- |
| **Demographics** | 34 (100%) | 12-42,50,55,63 |
| **Trust in authorities** | 17 (50%) | 12-14,16-18,20,23-26,28,31,34,36-38 |
| **Sources of news/information** | 19 (56%) | 12-16,18,21,23-26,30,31,34,36,39,41,50,55 |
| **COVID-19 Vaccine acceptance/intention proportion** | 32 (94%) | 12-29,31-36,38-42,50,55,63 |
| **Knowledge/attitude/ beliefs towards Covid-19 vaccine** | 26 (76%) | 12-17,19,20,22-28,31,33,35-40,42,55,63 |
| **Concern about COVID-19 vaccine safety/side effects** | 25 (74%) | 12-17,19,20,22-28,33,35-40,42,55,63 |
| **Concern about COVID-19 vaccine efficacy** | 17 (50%) | 12,14-16,22-26,33,37-40,42,55,63 |
| **Concern about COVID-19 vaccine development** | 7 (21%) | 13,14,16,17,20,38,39 |
| **Knowledge/attitude/behavior regarding COVID-19 disease** | 20 (59%) | 13,14,16-19,22,23,25,26,29,31,34,36,38-40,42,50,63 |
| **COVID-19 risk perception** | 15 (44%) | 13,14,16-19,22,23,25,26,34,36,38,40,63 |
| **History/ exposure to covid-19 in participants or their relations** | 16 (47%) | 13,15,17,19,20,22,24,25,27,28,30,31,33,34,36,39 |

**Table 4.** COVID-19 vaccine acceptance associated factors (n= number of studies examined the association between vaccine acceptance and each factor)

| **Study factors** |  | **Number of studies reported significant association (%)** | **Study ID** |
| --- | --- | --- | --- |
| **Sociodemographic factors** | **Age (n=28)** | 15 (54%) | 13-17,23-31,50,63 |
|  | **Gender (n=27)** | 12 (44%) | 12-22,32 |
|  | **Education (n=23)** | 13 (57%) | 12,13,15,17,18,20,21,23,27,31-33,55 |
|  | **Marital status (n=13)** | 4 (31%) | 12,14,27,35 |
| **Communication-related factors** | **Trust in authorities (n=8)** | 5 (63%) | 17,21,23,26,37 |
|  | **Sources of news/information (n=12)** | 7 (58%) | 12,18,26,30,31,41,50 |
| **COVID-19 Vaccine-related factors** | **Perceived vaccine safety/side effects (n=12)** | 10 (83%) | 14,19,22-25,36,39,40 |
|  | **Perceived vaccine efficacy (n=9)** | 8 (89%) | 14,22-24,38-40,42 |
|  | **Concerned about vaccine development (n=3)** | 3 (100%) | 14,17,39 |
| **Covid-19 infection-related factors** | **COVID-19 risk perception (n=10)** | 9 (90%) | 16,18,19,22,23,26,34,36,40 |
|  | **History/ exposure to covid-19 in participants or their relations (n=11)** | 3 (27%) | 17,20,36 |

### Sociodemographic factors

Most common sociodemographic factors associated with COVID-19 acceptance were gender, age, level of education, and marital status (Table 4). Female gender (11-21), younger age (14, 15, 17, 22-29), lower level of education (12, 16, 17, 19, 20, 22, 26, 30-32), and being single (11, 13, 26) were associated with lower vaccine acceptance in most studies.

Some of the other sociodemographic influencers of COVID-19 vaccine acceptance were sexual orientation (19),income (26, 33), region of origin (11, 17, 21), religion (11, 17), migratory background (20, 30, 34, 35), employment status (13, 20, 26), years living in host countries (22, 27, 32, 35, 36), racialized status (16, 31), working in health care settings (19, 24, 28), language fluency (27, 35), having comorbidities (29), and having insurance (31). In two studies, refugees living outside the refugee camps were less likely to accept COVID-19 vaccination (30, 37). Additionally, one study examined the relationship between different personality traits and COVID-19 vaccine hesitancy. In this study higher Conscientiousness, lower Openness and Neuroticism were associated with lower vaccine acceptance(20).

### Communication-related factors

Among the studies that examined the association between trust in authorities and COVID-19 vaccine acceptance, five studies (63%) found a positive association between level of trust in government and host country’s health care system and COVID-19 vaccine acceptance. Moreover, participants’ preference to get vaccinated against COVID-19 was significantly associated with their source of COVID-19 information in 58% of studies examining this association (table 4). Also, participants who had more exposure to confidence-inducing news and had no exposure to concerning news regarding COVID-19 vaccines were more likely to get the booster dose (22).

### COVID-19 Vaccine-related factors

Concerns regarding the safety/side effects of COVID-19 vaccine was a prominent factor associated with lower vaccine acceptability in 10 studies (83%) evaluating this association. Perceived vaccine efficacy was also another predictor of vaccine acceptability. Eight articles (89%) reported lower acceptance among participants who had mistrust in COVID-19 vaccines’ effectiveness. Another reason for lower vaccine acceptance was the belief that COVID-19 vaccines were developed too quickly and that the rushed pace of testing them would become problematic in the future. Being concerned about vaccine development was positively associated with vaccine hesitancy in all three studies evaluating this association. Additionally, six studies examined the participants’ knowledge/attitude scores towards COVID-19 vaccines (15, 22, 24, 34, 36, 38). Two studies showed that people with greater knowledge of COVID-19 vaccines had higher intention to get vaccinated (15, 22). Non-COVID vaccine refusal and negative attitude towards vaccines in general were associated with lower COVID-19 vaccine acceptance (11, 29, 39). Also, getting the seasonal flu vaccination (13, 38) were a positive predictor of vaccine acceptance.

### COVID-19 infection-related factors

Among studies that examined the association of COVID-19 risk perception and vaccine acceptability, nine (90%) found a positive association; whereby, participants who had higher COVID-19 risk perception had higher vaccine acceptability. Only three out of the 11 studies that evaluated the relationship between COVID-19 vaccine acceptance and previous history of exposure/infection with COVID-19 disease reported a significant association. In the study by Frisco et al. previous infection with COVID-19 was associated with higher vaccine acceptance (16), while Ogunbajo et al. suggested that previous positive COVID-19 test was higher among vaccine hesitant participants (19). Moreover, West et al. found that vaccine acceptance was higher among workers who had more COVID-19 exposure through crowded housing (35). Also, Rego et. al did not find any association between knowing someone with COVID-19 infection and vaccine acceptance (30). Four studies examined the association of vaccine acceptability with knowledge and attitudes regarding COVID-19 infection. All reported that higher COVID-19 infection knowledge and attitude scores were associated with higher vaccine acceptability (13, 24, 33, 38). Participants’ health behaviors during COVID-19 pandemic were other factors measured in some studies. They found wearing a face mask and social distancing were associated with higher COVID-19 vaccine acceptance/intention (24, 30, 33).

### Other factors associated with COVID-19 vaccine hesitancy

Fear of needles (21, 39), belief in house remedies (39), religious reasons (23, 33, 39), belief in natural exposure to germs and viruses as the best protection and “not to mess up with nature” (13, 39) were other reasons for lower vaccine acceptance as reported by vaccine hesitant or anti vaccine individuals.

Shaw et al. reported that individuals with high vaccine acceptability had a lower social vulnerability index(40). Another study by page et al. showed that higher immigration enforcement exposure, such as experiencing or knowing someone who experienced immigration raids, detention or deportation, was associated with lower odds of COVID-19 vaccine acceptance (29). In contrast, acculturation and social integration of migrants were associated with higher vaccine acceptance (32, 36), as well as cues to action (15, 41) and social norms (15). Cues to action, refers to being motivated to get vaccinated by seeing neighbors, community leaders, doctors, or politicians receive the vaccine; whereas vaccination as a social norm refers to the higher likelihood of getting vaccinated when most people an individual knew had received the vaccine.

## Discussion

Our findings suggest that predictors of COVID-19 vaccine acceptance among migrant populations can be categorized in four major groups: “sociodemographic factors”, “communication-related factors”, “COVID-19 vaccine-related factors” and “COVID-19 infection-related factors”. Lower vaccine acceptance was associated with mistrust in the host countries’ government and healthcare system, concerns about the safety and effectiveness of COVID-19 vaccines, limited knowledge of COVID-19 infection and vaccines, lower COVID-19 risk perception, and lower integration level in the host country. Also, female gender, younger age, lower education level, and being single were sociodemographic factors associated with lower vaccine acceptance in most studies. Sources of information regarding COVID-19 and vaccines and previous history of COVID-19 infection were other influencers of vaccine acceptance.

A systematic review on qualitative studies on COVID-19 vaccine hesitancy among ethnic minorities similarly identified five major themes for drivers of vaccine hesitancy among these populations: 1) Institutional mistrust, 2) Lack of confidence in vaccine and development process, 3) Lack of reliable information or messengers, 4) Complacency/perceived lack of need and 5) Structural barriers to vaccine (42).

While our findings show that most studies reported lower vaccine acceptance among immigrant women, as the association between sex/gender and COVID-19 vaccine acceptance among RIM was significant in only 44% of studies examining this association, further analytic investigation is needed on this matter. In a meta-analysis conducted by Alimoradi et al. the pooled prevalence of Covid-19 vaccine acceptance did not significantly differ according to gender in migrants and ethnic minorities (43). Another systematic review on determinants of routine and COVID-19 vaccine uptake among RIM also reported no strong association with gender (44). Higher skepticism among females might be even more important to achieve higher vaccination coverage, as this can directly affect children COVID-19 vaccination rate, because mothers appear less willing to vaccinate their children against COVID-19 than fathers (45, 46). Understanding the sociodemographic predictors of vaccine acceptability can help to improve community engagement strategies to increase vaccination uptake in populations with lower vaccine acceptance, such as outreach initiatives for under-immunized ethnic groups. One example is a successful program addressed low vaccination rates among women of Rohingya refugees in Cox’s Bazar, Bangladesh. Strategies included educational programs, women-only radio clubs, religious group-study sessions, and employing female vaccinators. These measures improved access to accurate information and created a comfortable environment for refugee women to get vaccinated (47).

Other sociodemographic factors of potentially great importance among migrant populations include migration status, years living in the host country, region of origin, language fluency, religion, and immigration enforcement exposure (11, 17, 21, 27, 32, 34, 48). Evidence suggests that language barriers and cultural variations are prominent factors that can affect these populations’ intention to be immunized (11).

The term “vaccine hesitancy” might imprecisely blame individuals, while there are several fundamental factors that may lead to making this decision (49). Our findings corroborate previous research that suggests trust in the host country’s government and public health authorities are key factors that affect migrants’ attitude towards COVID-19 vaccines and vaccination decision making (22, 30, 36, 49, 50). Lack of trust among migrant communities can stem from previous experience of xenophobia, racial discrimination, and anti-migrant politics (50, 51). A qualitative systematic review of COVID-19 vaccine hesitancy by Shearn et al. showed that “feeling unheard, ignored or excluded from the healthcare system” caused institutional mistrust resulting in COVID-19 vaccine hesitancy among RIM populations (42). Moreover, trust also plays a crucial role in acceptance of non-COVID vaccines among RIM population (50).

Previous research also suggests that source of information is an important influencer of vaccination decision making (52); however, this factor might be more complicated among RIM populations based on different cultural and linguistic backgrounds and adaptation level within host countries. Moreover, the lack of adequate information in migrants’ primary languages often lead to reliance on community networks, traditional and social media, which might lead to confusion and inability to discern factual information from misinformation (25, 29, 53). Similarly, in non-COVID vaccination context, the overwhelming amount of contradictory information and false claims spread through social media regarding vaccines undermines the efforts to encourage vaccine acceptance among migrants (50).

A systematic review by Romate et al. suggest that the perceived safety and effectiveness of COVID-19 vaccines as well as risk perception of COVID-19 infection is associated with both “knowledge” and “trust” factors (54). Additionally, our study revealed that increased knowledge level about both COVID-19 infection and vaccines among migrants were associated with higher vaccine acceptability. These findings reinforce previous research that investigated these associations among general populations (55, 56). Applying various community engagement strategies may help build trust and address knowledge gaps and misinformation among these diverse communities (57). Previous research suggests “using high-touch rather than high-tech approaches” and community health workers’ involvement help provide trustworthy sources of information and tailoring new health recommendations (49).

Finally, our study suggests that higher levels of migrants’ acculturation and integration in the host country is associated with higher COVID-19 vaccine acceptance (32, 35, 36). Previous research similarly showed that migrants living in societies with lower integration policies for migrant populations, experience poorer health conditions (32, 58, 59) Strategies that facilitate migrants’ psychological, social, economic, political, navigational, and linguistic integration in a host country can, in turn affect other vaccine hesitancy related factors such as trust in authorities and misinformation (32).

### Implication for public health Policy

Our findings may help inform the programs and community outreach strategies to improve uptake of COVID-19 vaccines in migrant and refugee subgroups with lower vaccine acceptance. Reducing mistrust in authorities and addressing knowledge gaps among migrant populations is crucial for improving COVID-19 vaccine acceptance and similar public health challenges.

## Limitations

It is noteworthy that we noticed some limitations in the included studies. Due to the cross-sectional design of included studies, we cannot make causal claims. Most studies used non-random sampling strategies, which meant that participants might not have been good representatives of migrant populations. Further, using online platforms to recruit study participants and survey in English or the host country’s language, can skew the data towards more acculturated, connected, and less vulnerable study participants. Moreover, most studies did not consider potential confounding variables such as having comorbidities, health literacy, duration of residence in host country in their analyses. Finally, recall bias, selection bias, and social desirability bias were other issues which might have affected the included studies’ findings.

## Future studies

Evaluation of migrant population’s attitude towards the COVID-19 booster dose(s) and different types and brands of COVID-19 vaccines would be beneficial. Undertaking a systematic review with an equity lens might be helpful to better identify subpopulations of migrants with suboptimal vaccination uptake.

## Conclusion

The acceptability of COVID-19 vaccines among RIM is influenced by various factors, including sociodemographic characteristics, communication-related factors, COVID-19 vaccine-related factors, and COVID-19 infection-related factors. Targeted vaccination plans, community engagement strategies, and efforts to address knowledge gaps and build trust are crucial for promoting vaccine acceptance among RIM populations.

## Data Availability

All relevant data are within the manuscript and its Supporting Information files.

## Acknowledgment

This research was funded by the Canadian Institutes of Health Research (CIHR).

All authors were actively involved in discussing and refining the research questions. Kevin Pottie provided expertise in designing research objectives, defining the inclusion and exclusion criteria and search strategy as well as manuscript revision. Gabriel Fabreau and Denise Spitzer contributed to formulating the research objectives and editing the manuscript, ensuring its quality and accuracy throughout the process. Yasaman Yazdani and Poojitha Pai contributed to defining the search strategy, conducting the literature search, and screening articles for inclusion, data extraction, synthesizing the findings, and drafting sections of the report. Shahab Sayfi contributed to the screening process and interpretation of the results. Arash Mohammadi and Saber Perdes contributed to manuscript revision. All authors reviewed and approved the final manuscript for publication. The authors have no conflicts of interest to declare.

We would like to thank Ms. Roxanne Isard, research and scholarly communication librarian at Western University, for her helpful guidance in developing this study’s search strategy.

## Appendix 1. Medline search strategy

## Appendix2. Data extraction tool

